# Clinical practice recommendations on lipoprotein apheresis for children with homozygous familial hypercholesterolemia: an expert consensus statement from ERKNet and ESPN

**DOI:** 10.1101/2023.11.14.23298547

**Authors:** M. Doortje Reijman, D. Meeike Kusters, Jaap W. Groothoff, Klaus Arbeiter, Eldad J. Dann, Lotte M. de Boer, Sarah D. de Ferranti, Antonio Gallo, Susanne Greber-Platzer, Jacob Hartz, Lisa C. Hudgins, Daiana Ibarretxe, Meral Kayikcioglu, Reinhard Klingel, Genovefa D. Kolovou, Jun Oh, R. Nils Planken, Claudia Stefanutti, Christina Taylan, Albert Wiegman, Claus Peter Schmitt

**Author notes:** **Corresponding author:** Albert Wiegman, Amsterdam UMC, location AMC Department of Paediatrics, Meibergdreef 9, 1105 AZ Amsterdam, The Netherlands. These authors contributed equally (Albert Wiegman, Claus Peter Schmitt).

## Abstract

Homozygous familial hypercholesterolaemia is a life-threatening genetic condition, which causes extremely elevated LDL-C levels and atherosclerotic cardiovascular disease very early in life. It is vital to start effective lipid- lowering treatment from diagnosis onwards. Even with dietary and current multimodal pharmaceutical lipid- lowering therapies, LDL-C treatment goals cannot be achieved in many children. Lipoprotein apheresis is an extracorporeal lipid-lowering treatment, which is well established since three decades, lowering serum LDL-C levels by more than 70% per session. Data on the use of lipoprotein apheresis in children with homozygous familial hypercholesterolaemia mainly consists of case-reports and case-series, precluding strong evidence-based guidelines. We present a consensus statement on lipoprotein apheresis in children based on the current available evidence and opinions from experts in lipoprotein apheresis from over the world. It comprises practical statements regarding the indication, methods, treatment targets and follow-up of lipoprotein apheresis in children with homozygous familial hypercholesterolaemia and on the role of lipoprotein(a) and liver transplantation.

## Introduction

Homozygous (both true and compound heterozygous) familial hypercholesterolaemia (HoFH) is a rare disease characterized by extremely elevated low-density lipoprotein cholesterol (LDL-C).^1^ These elevated LDL-C levels, present from birth, may cause life-threatening atherosclerotic cardiovascular disease (ASCVD) early in life, if not treated sufficiently. To prevent this, it is vital to diagnose HoFH as early as possible and start treatment from diagnosis onwards.^1^ For many HoFH patients, it is mandatory to combine the optimal use of the currently available pharmacological lipid-lowering therapies (LLT) with lipoprotein apheresis (LA).^2^

LA comprises several methods of selective therapeutic apheresis which reduce LDL-C levels by more than 70% per session.^3, 4^ LA has to be repeated weekly or biweekly, as LDL-C levels increase after each session.^3, 5^ Although the lipid-lowering effect of LA is strong, its impact on preventing ASCVD in HoFH is difficult to analyse. HoFH is rare, LA is not available for many patients around the globe and if available, it is not ethical to withhold children from LA to analyse the clinical efficacy. From historical reports on HoFH patients, we know that without LA treatment, severe life-threatening ASCVD may occur in early childhood.^6^

There is an unfulfilled need for guidance on the treatment of LA in children with HoFH, especially in the current times with major advances in pharmacological LLTs.^7^ Therefore, this consensus statement, initiated by the European Rare Kidney disease Network (ERKNet) and endorsed by the European Society of Paediatric Nephrology (ESPN) Dialysis Working Group, was developed to provide guidance to healthcare professionals on the treatment of LA in children with HoFH. These recommendations are based on the available evidence from observational studies in children and adults and the opinion of experts in the field of HoFH. Because the rareness of the disease and ethical considerations, there are no RCTs or prospective interventional studies on the LDL-C lowering effects of LA, and therefore the quality of evidence for many statements remains low.^7^ Hence, it is important to use these statements as a guidance and adapt them to the individual patient’s needs. In this consensus statement, we propose statements on the use of LA in the treatment of children with HoFH, including indication, methods, vascular access, treatment target, monitoring clinical efficacy, side effects and the role of Lp(a) and liver transplantation.

## Methods

### The consensus statement development groups

A core workgroup and a voting group were involved in the consensus statement. Participants were recruited via ERKNet and two international HoFH registries: HICC (HoFH, the International Clinical Collaborators; NCT04815005) and CHAIN (Children with Homozygous hypercholesterolemia on lipoprotein Apheresis: an International registry). The core workgroup involved 21 HoFH experts from ten countries and working in different medical specialties (Supplementary table 1). The voting group consisted of 19 experts in HoFH with different medical specialties from 13 countries (Supplementary table 2). The core workgroup formulated key questions, performed a literature review, wrote the statements and rationales, rated the quality of evidence and wrote the manuscript. Subgroups worked on the statements and rationales on indication, methods of LA, vascular access, treatment target, monitoring clinical efficacy, monitoring side effects and the role of Lp(a) and liver transplantation. The voting group was asked to share their level of agreement and feedback for each statement.

### Developing clinical questions

To give specific recommendations, clinical questions were developed for each topic as a basis for creating statements. These clinical questions were based on an overall question including patients, intervention, comparator and outcome (PICO).^8^ Patients were children under the age of 18 with a genetic or clinical diagnosis of HoFH following the criteria from Cuchel et al.^1^ HoFH can be diagnosed genetically by confirmation of two pathogenic variants in the *LDLR*, *APOB, PCSK9*, or *LDLRAP1* gene, or can be diagnosed clinically when a patient has untreated LDL-C levels >10 mmol/L (>∼400 mg/dL) together with either cutaneous or tendon xanthoma before the age of 10 years or untreated LDL-C levels consistent with HeFH in both parents.^1^ The intervention was treatment with LA and the comparator was standard of care without LA. The outcome was safety, and efficacy in terms of LDL-C lowering, clinical and imaging findings of ASCVD. ASCVD was defined as ASCVD related to HoFH including angina pectoris, aortic stenosis, myocardial infarction, sudden cardiac death, percutaneous coronary intervention (PCI), coronary artery bypass graft surgery (CABG) and aortic valve replacement.

### Literature review and studies included

PubMed and Embase databases were searched until 22^nd^ of November 2021. All articles in English describing paediatric patients with HoFH were selected irrespective of the design of the study. For each topic, articles including relevant information for the specific topic were selected. To include the most recent literature up to April 2023, members of the workgroup added 11 additional references during the writing process, which were published after the database searches were performed. If this resulted in a need for adjustment of a statement, this was again reviewed by all participants. This occurred once, based on the most recent guidelines for LDL- cholesterol targets.^1^

### Grading system

We applied the grading system from the American Academy of Pediatrics (AAP) (Figure 1).^9^ For each statement, the quality of evidence was graded and the strength of the recommendation was chosen based on the assessment of benefit or harm.^9^

After the core workgroup agreed on the content and grading of the statements, these were sent to an external voting group, which was asked to share their level of agreement on a five-point Likert scale (strongly disagree, disagree, neither agree/disagree, agree and strongly agree) for each statement. Consensus was regarded as a minimum of 70% of the voters choosing ‘agree’ or ‘strongly disagree’ for each statement. If this was not reached, the statement was either revised or removed by the core working group based on the suggestions of the voting group, and proposed at the voting group again, until 70% agreement was reached. Finally, 70% agreement was obtained for 22 out of 24 statements. Two were removed as statement and brought as topics of discussion within the rationales on the specific topics.

## Consensus Statements

### A. Indication for LA

#### Statements

1. We recommend starting LA in children diagnosed with HoFH if LDL-C levels are >7.8 mmol/L (>300 mg/dL) despite optimal lipid-lowering therapy. (X - strong)
2. We recommend starting LA in children diagnosed with HoFH <u>and (subclinical) ASCVD</u> if LDL-C levels are >3.4 mmol/L (>130 mg/dL) despite optimal lipid-lowering therapy. (X - strong)
3. We suggest considering starting LA in children diagnosed with HoFH <u>without (subclinical) ASCVD</u> if LDL-C levels are between 3.4 mmol/L (130 mg/dL) and 7.8 mmol/L (300 mg/dL) despite optimal lipid- lowering therapy. (X - moderate)
4. We recommend starting LA early as possible in life. (X - strong)

#### Rationale

*Threshold of LDL-C level for LA indication.* The fundamental rationale for anticipating LA treatment in HoFH children resides in the reduction of very-high LDL-C exposure that is associated with significant ASCVD risk.

There is sufficient evidence, that for prevention of ASCVD, the paradigm for LDL-C target level is, “the lower the better”. Current recommendations are that ideal LDL-C levels should be below 1.8 or 2.5 mmol/L (70 or 100 mg/dL) for the adult population without signs of ASCVD.^1, 10^ For HoFH patients, these targets are difficult to reach under the current therapeutic options. The *threshold* of LDL-C for the indication of LA has originally been established on >13 mmol/L (>500 mg/dL) for HoFH and >7.8 mmol/L (>300 mg/dL) for heterozygous familial hypercholesterolemia (HeFH) by the FDA.^11^ We follow the recommendation of both the American Heart Association and the National Institute for Health and Care Excellence to start LA in both HoFH and HeFH patients with LDL-C levels >7.8 mmol/L (>300 mg/dL) despite optimal LLT. Optimal LLT is regarded as diet combined with multimodal pharmacological LLT. This can include ezetimibe, statins, proprotein convertase subtilisin/kexin type 9 (PCSK9) inhibitors, evinacumab or lomitapide, depending on which LLTs are available, affordable and tolerable for the patient.^7^ There is consensus that the threshold for LA should be lower in FH patients with established ASCVD. In Germany, a threshold for all FH patients with ASCVD of 3.4 mmol/L (130 mg/dL) is recommended, which is generally regarded as the threshold of high-normal LDL-C level.^12^ Therefore, we recommend starting LA in children diagnosed with HoFH and (subclinical) ASCVD if LDL-C levels are >3.4 mmol/L (>130 mg/dL) despite optimal LLT. If LDL-C levels are between 3.4 mmol/L (130 mg/dL) and 7.8 mmol/L (300 mg/dL) despite optimal lipid-lowering therapy and no (subclinical) ASCVD is present, we suggest considering starting LA based on individual circumstances, such as age, vascular access options and additional cardiovascular risk factors.

*The age at LA treatment commencement* is associated with ASCVD event-free survival, together with treatment duration, and the current on-treatment LDL-C levels.^13^ In theory, the same paradigm would be applicable as in the threshold discussion, “the sooner, the better”. LA is technically feasible in very young children and has been successfully chronically executed in children from the age of 2.3 years, provided there is a trained team that can face technical limitations such as low blood flows, consequence of the use of small calibre needles, and the risk of mild hypotension related to relatively high extra-corporal volumes.^3, 14–17^ Observational studies support early onset of LA therapy in HoFH.^16, 18–22^ LA seems more effective in preventing development of ASCVD than stopping further deterioration of already acquired ASCVD, which would be an extra argument for early onset of therapy.^19^

### B. Methods of lipoprotein apheresis

#### Statements

5. We suggest using selective methods for lipoprotein apheresis, for which equally efficient systems are available. (C – moderate)
6. We suggest aiming for an acute reduction of LDL-C plasma levels of at least 70% with each apheresis session. (X – moderate)

#### Rationale

LDL-C can be removed by unselective plasma exchange and by selective LA methods, either as a plasma separation method or as a whole blood method.^23^ Currently, there are six technically different options for selective extracorporeal LA available (see table 1 for comparison).^24, 25^

**Table 1.** Comparison of lipoprotein apheresis methods.

| Method | Extra corporeal volume | Selectivity | HDL-C removal | Fibrinogen removal | Lp(a) removal | Reported side effects | Advantages | Disadvantages |
| --- | --- | --- | --- | --- | --- | --- | --- | --- |
| **Polyacrylate full blood adsorption (DALI, Fresenius)**<br>*haemoperfusion – whole blood technique* | 330-705 cc<sup>a</sup> | ++ | +/- | - | ++ | Bradykinine-related<sup>3</sup> | Very effective, selective, simple | Expensive, high extra-corporeal volume, contra-indication ACEi |
| **Kaneka LA 15**<br>*Dextran sulphate adsorption - plasma separation* | 130-160 cc<sup>b</sup> (300 cc total) | ++ | +/- | - | ++ | Allergic, Bradykinine-related<sup>3</sup>, hypoCa | Very effective, selective, relatively low extra corporal volume | Complicated technique, contra-indication ACEi |
| **Dextran sulphate full blood adsorption (Liposorber D, Kaneka)**<br>*Haemoperfusion - whole blood technique* | 394 cc (DL-50), 484 cc (DL-75), 690 cc (DL-100) | ++ | +/- | - | ++ | Bradykinine-related<sup>3</sup>, hypoCa | Very effective, selective, simple | Expensive, high extra-corporeal volume, contra-indication ACEi |
| **HELP (Braun)**<br>*heparin-induced LDL precipitation* | 150 cc<sup>b</sup> (450 cc total) | +<sup>5</sup> | + | ++ | ++ | Potentially bleeding risk, low blood pressure | Safe, proven impact on outcome, anti-inflammatory | Less selective, less effective, loss C3/C4, high loss of fibrinogen, complicated technique |
| **Double/cascade filtration (DFPP)**<br>*Plasma separation and filtration* | 222 cc | + | ++ | + | ++ | Occasionally hypotension, nausea, vomiting | No bradykinin release, simple technique | Less selective, less effective on the long run, loss of IgG, Alpha-2-Macroglobulin |
| **Immuno-adsorption**<br>*Anti Apolipoprotein B (Therasorb)* | 180 cc<sup>b</sup> | + | +(+) | +(+) | ++ | Occasionally hypotension, nausea, vomiting | Effective, selective | less effective and selective than other selective methods, expensive, relative long procedure |
| **Plasma exchange**<br>*Plasma filtration or plasmapheresis (centrifuge)* | 140-185 cc | - | ++ | ++ | ++ | Bleeding risk, fibrinogen loss, hypotension, | Most simple technique, widely used, cheap | Unselective, less effective, loss of plasma proteins, HDL-C removal, bleeding risk, anaemia |

<sup>a</sup>Depending on filter size 500, 750, 1000 & 1250 cc; <sup>b</sup>expressed as estimated blood equivalent volume for the blood separation systems (LA15, HELP, immuno-adsorption; total = blood and plasma volume); <sup>c</sup>abdominal pain, flushing, hypotension.
5 Data from Bambauer 2012, Klingel 2003 and information from the companies.<sup>24, 25</sup>
Abbreviations: ACEi, ACE-inhibitor; HELP, Heparin Induced LDL precipitation; HDL-C, high-density lipoprotein cholesterol; hypoCa, hypocalcaemia; LA, lipoprotein apheresis; LDL-C, low-density lipoprotein cholesterol; Lp(a), lipoprotein(a);

Although all existing systems are effective in removing LDL-C, a review of 76 studies on LA in FH, showed that in daily practice, selective methods (LA15, polyacrylate full blood adsorption and dextran sulphate full blood adsorption) were slightly more effective than plasma exchange (on average 71 vs. 63% LDL-C removal).^20^ Within the selective techniques, there are small differences in efficacy.^26–28^ Apart from slightly less efficient LDL-C removal, plasma exchange and double filtration plasmapheresis (DFPP) bare the problem of removal of other components, such as HDL-C, fibrinogen and IgG.^29, 30^

**Experience of the authors:** most data on efficacy are derived from studies in adults. Contrary to often lower reported values in adult patients, in children >70% LDL- C reduction per session can be achieved with selective techniques like HELP, LA15 and DFPP, as long as enough plasma is exchanged, since these systems do not saturate in children. Limitation of LA efficacy in daily practice with children is often caused by LA duration with limited blood and plasma flow due to poor vascular access quality. The recommended plasma exchange volume for the plasma separation techniques are between 40 and 60 cc/kg; for the whole blood systems between 1.3 and 1.5 times the total blood volume.

*LA in small children*: The extracorporeal volume can limit the use of some techniques in small children. This accounts especially for the whole blood systems. Most plasma separations techniques have been applied successfully in children aged >2 years old, with most experience in young children has been achieved with the LA15 Kaneka system that has a blood equivalent volume of 130 cc.^15^ Patients >13 kg do not need blood priming with this system. Practical advice for the management of LA in young children includes a low blood flow at onset of therapy, starting with 15-30 ml/min, priming the extracorporeal system with normal saline or, in case of low blood pressure, human albumin and the involvement of skilled nurses experienced in conducting extra-corporeal blood purification techniques in children.

In conclusion, all available selective methods lead to a significant removal of LDL-C with preservation of other proteins. Acute reductions >70% are achievable, as long as optimal plasma volumes per kilogram body weight are applied.^3^ Therefore, we suggest aiming for an acute LDL-C reduction of at least 70% per LA session.

### C. Vascular access

#### Statements

7. Vascular access options should be discussed with the patient and families in detail and individual decisions should be taken considering age, vascular anatomy and individual needs. (ungraded)
8. In children not suited for peripheral vein puncture, we suggest starting with a functioning AVF. (C - moderate)
9. In children with AVF, the cardiac burden of the increased circulating volume should be monitored. (D - weak)
10. Vascular access should be placed and used by well-trained personnel. (ungraded)

#### Rationale

Various forms of vascular access are applicable for LA in youth: 1) peripheral vein needling; 2) arteriovenous fistula (AVF) or arteriovenous graft; and 3) a tunnelled central venous line (CVL), with or without a port. The provision of the optimal vascular access for LA should be patient focused, and based on a multidisciplinary approach in assessing and educating patients. Complications of vascular access depend on the access type (Table 2). Specific advantages and disadvantages of the different vascular access options should be explained, such as the need of regular needling and associated pain, the option of local anaesthetic crèmes to reduce puncture pain, LA session duration, and the associated risks of infection and access dysfunction due to dislocation and clotting, eventually requiring novel access placement at a different site.

**Table 2.** Comparison of vascular access types.

| Peripheral vein needling |  | Arteriovenous fistula |  | Central venous line |  |
| --- | --- | --- | --- | --- | --- |
| Pro | Cons | Pro | Cons | Pro | Cons |
| Ready to use | Puncture pain | Less stigmatization than with CVL | Puncture pain | Pain free | Stigmatization |
| No stigmatization, normal body perception | Difficulties with frequent needling | Highest blood flow, short treatment time | Vascular surgery required | Safe vascular access for frequent use | High infection and obstruction (clotting) rates |
| No impact on physical activities | Low blood flow, longer session duration | Very low infection rate, lower rate of obstruction (clotting) than CVL | Difficult in small children, and need of long maturation time | Feasible in small children | Risk of dislocation |
|  |  | Safe vascular access for frequent use | Potential cardiac burden with high shunt volume |  | Risk of thrombi/emboli and central vascular stenosis |
|  |  | Less interference with physical activities than CVL |  |  | Interferes with physical activities |

3 Abbreviation: CVL, central venous line.

**Table 3.** Most commonly described potential adverse effects of lipoprotein apheresis.

|  |  |
| --- | --- |
| **Clinical symptoms** | Hypotension |
|  | Nausea/vomiting |
|  | Stomach ache |
|  | Fatigue |
|  | Dizziness |
|  | Angina |
|  | Anaphylactic reactions (cutaneous flushing, nausea, headache and hypotension) |
|  | Tingling/urticaria |
| **Bradykinin release related** | Nausea and vomiting, flushing, tongue swelling, severe abdominal pain, hypotension |
| **Iron deficiency** | Need for iron supplementation |
| **Vascular access related** | Puncture difficulties |
|  | Insufficient blood flow |
| **Lipoprotein apheresis related** | Obturation of the blood lines |

Observational evidence suggests peripheral vein access for LA is feasible in children, with safe puncturing of the veins and pain tolerated.^18^ Achievement of therapeutic targets, number of missed sessions and the individual burden of regular vein punctures require close monitoring. In all other children, a permanent vascular access for long-term treatment needs to be established.

There is limited literature available on the use of AVF and CVL in children with HoFH. Even though concerning a different patient population, studies in children on chronic haemodialysis show that AVF is to prefer over CVL, as CVLs are associated with a significantly higher risk of infection, access dysfunction, access replacement and vascular stenosis than AVF.^14, 16, 31–34^ The duration of the LA sessions is shorter with AVF, and a non-significant trend towards lower mean LDL-C levels has been observed.^3^ It is unclear to what extent arteriovenous grafts are an alternative in children with veins too small for AVF.

In AVF, blood flows from the high resistance arterial system into the low resistance venous system, with a subsequent rise in venous return and cardiac output. It decreases arterial impedance and thus lessens the left ventricular afterload. The lowering of arterial impendence may also reduce the effective circulating volume of the systemic circulation, activating arterial baroreceptors, and leading to secondary increase in cardiac sympathetic tone, contractility, and output.^35–37^ The impact of these effects of AVF on the cardiac function is controversial.^38^ In patients on haemodialysis, the vast majority of patients tolerate AVF well.^39^ At present, it is unknown in how far the AVF associated cardiac burden due to the increased circulating volume may represent an additional cardiac risk in children on LA, respective AVF shunt volume should be monitored and AVF with large shunt volumes may require surgical flow reduction.

Vascular access sites are limited. Improved outcomes have been reported when skilled surgeons work with dedicated vascular access clinics.^40^ Preoperative diagnostics and site selection, aseptic technique for access use, vascular access monitoring and prevention of thrombosis have recently been described in a consensus document for children requiring maintenance haemodialysis by the European Society for Paediatric Nephrology Dialysis Working Group; these recommendations apply for children on LA with an AVF as well.^41^ Recent studies suggest that AVF, if provided by specialized surgeons, can be placed in children aged less than 2 years.

Complications are thrombosis, stenosis or non-maturation occur in a minority, but maturation times of up to 6 months have been reported.^34, 42, 43^

**Experience of the authors:** five of the 10 centers of the core workgroup performing LA in children have positive long-term experience with peripheral vein puncture in children from the age of 3-6 years onwards with weekly to monthly LA and session durations of 2-6 hours. Five centers primarily use AVF in children from 3 to 12 years onwards with weekly to monthly sessions as needed and session durations of 1.25-3 hours. Successful 12 years usage of arteriovenous graft (AVG) was reported in a patient with veins too small for AVF creation. CVL were used in one center in children aged 3-7, but with high complication rates.^14^ One center reported routine use of ports from the age of 6 years onwards, with weekly to twice-monthly sessions and 4-8 hours of session duration; one center used a port for temporary vascular access in one patient. Due to the long treatment times and the need for surgical replacement of the port every few years, this vascular access may not be a preferable option. Most centers had a strong preference for one of the vascular access options, mainly related to the specialty of the treating physician. Nephrologists most often used AVFs whereas cardiologists used peripheral veins.

### D. Treatment target

#### Statements

11. In between weekly or biweekly apheresis sessions, we suggest using the adjusted Kroon formula to calculate the mean LDL-C plasma levels: LDL-Cmean = LDL-Cpost + K (LDL-Cpre – LDL-Cpost). LDL-Cpost: LDL-C level directly after the LA session, LDL-Cpre: LDL-C level directly before the LA session. K: rebound coefficient, 0.65 for HoFH patients. (C – weak)

12. We recommend aiming for a mean LDL-C treatment target of <3.0 mmol/L (<115 mg/dL). (X – strong)
13. For children with ASCVD, we suggest considering lower mean LDL-C treatment targets of <1.8 mmol/L (<70 mg/dL). (X – moderate)
14. We suggest considering reducing the frequency of lipoprotein apheresis if the mean LDL-C plasma levels stay within the treatment target with the use of newly available effective lipid lowering drugs. (X – moderate)

#### Rationale

Although high-level evidence exists on the benefits of lowering serum LDL-C concentrations with respect to reducing cardiovascular risk,^44, 45^ there is no evidence for the optimal LDL-C target for HoFH children on LA. Previously, the proposed treatment target for children with HoFH was set on <3.4 mmol/L (<130 mg/dL).^46^ However, observational studies have shown early development of ASCVD in children reaching these treatment targets.^16, 20^ Recently, it was suggested to further lower the treatment target in children with HoFH to <3.0 mmol/L (<115 mg/dL).^1^ In line with this recommendation, we recommend aiming for a mean LDL-C treatment target of <3.0 (<115 mg/dL) in children with HoFH on LA. For children with (subclinical) ASCVD, we suggest considering lower LDL-C targets of <1.8 mmol/L (<70mg/dL), in line with the previous suggested treatment target for adults with HoFH and ASCVD.^46^ We believe that a lower target may be too challenging and burdensome for children in daily practice and consequently not feasible in children with the current available therapeutic options.

Another reason to support these targets is the introduction of new, highly effective lipid lowering agents. The PCSK9 antibody evolocumab reduced LDL-C by 45% in an RCT including 157 paediatric HeFH patients and reduced LDL-C by 30% in two children with HoFH.^47, 48^ Evinacumab, a monoclonal antibody against ANGPTL3, reduced LDL-C by almost 50% on top of background LLT in 65 HoFH patients, including one adolescent.^49^ In a recent observational report, two paediatric HoFH patients (12 and 16 years of age) were treated with a statin, ezetimibe and weekly LA. Addition of evinacumab decreased mean pre-apheresis LDL-C levels from 5.5 and 5.1 mmol/L to 2.5 and 2.2 mmol/ L, respectively. Total plaque volumes were reduced by 76% and 85% after 6 months of evinacumab treatment^50^, demonstrating that with drastic LDL-C lowering, atherosclerotic plaques can regress at young age, even in HoFH patients. With the accomplishments of the novel lipid-lowering therapies it might also be safe to reduce the frequency of LA, provided the mean LDL-C plasma levels stay within treatment targets. However, their efficacy in the prevention of ASCVD and mortality in paediatric patients has yet to be proven.

*How to monitor LDL-C:* while with lipid-lowering drugs, the LDL-C levels are relatively stable over time, with LA treatment, the LDL-C levels have a saw tooth-like pattern. Pre-LA LDL-C levels give an overestimation of the actual LDL-C level over time, while post-LA LDL-C levels give a considerable underestimation. Time-averaged concentrations provide the best estimate of the mean LDL-C levels over time.^51^

The Kroon formula was developed to estimate the mean LDL-C levels between two LA sessions on a biweekly scheme and based on a study on the rebound of lipoproteins in 20 hypercholesterolaemic (no HoFH) adult men after LA with a rebound coefficient of 0.73.^51^ Thompson et al.^52^ calculated a rebound coefficient specifically for HoFH patients of K=0.65 based on 11 adult HoFH patients who received biweekly LA. No large validation studies have been performed to evaluate this formula and no studies have been performed to validate this coefficient in paediatric patients nor in patients receiving LA at different frequencies other than biweekly. Therefore, the suggested Kroon formula with a coefficient of 0.65 only provides a rough estimate of the mean LDL-C levels in paediatric HoFH patients on LA once every two weeks and might even be less reliable if other frequencies are applied.

### E. Monitoring clinical efficacy

#### Statements

15. We recommend performing echocardiography (with colour and Doppler) annually (X - strong)
16. We recommend performing low dose CCTA before LA therapy is initiated (X - strong)
17. We recommend, if potentially obstructive atherosclerotic plaque >50% is visible on CTCA, to detect and evaluate potential coronary ischemia by non-invasive functional test (X - strong)
18. We recommend cardiac catheterization and coronary angiography for HoFH children when invasive intervention may be required, and for coronary imaging if CCTA is not feasible (X - strong)

#### Rationale

Asymptomatic children with HoFH should be screened for subclinical ASCVD. Common consequences of HoFH include aortic stenosis (AS) and coronary non-calcified or calcified plaques.^46^ AS is mainly evaluated by echocardiography, and seldom by invasive cardiac catheterization, contrast angiography or cardiac magnetic resonance imaging (MRI).^53, 54^ Presence of (non-) calcified plaques, perivascular inflammation and hemodynamic obstructive lesions can be evaluated by prospective ECG-triggered CT coronary angiography (CTCA). Potential ischemia due to obstructive coronary artery stenoses should be primarily evaluated by non-invasive functional tests, such as exercise ECG, stress perfusion with PET-CT, cardiac MRI or Single Photon Emission CT Myocardial Perfusion (SPECT).

<u>Ultrasonography:</u> subclinical and clinical aortic valvular disease (VD), ostia and proximal segments of the coronary arteries (not always easy to visualize), and any abnormality in cardiac function in HoFH children can be assessed by echocardiography with colour and Doppler.^55^ Aortic valve regurgitation or stenosis, aortic root thickening, narrowing of the ostia and proximal segments of the coronary arteries, and abnormal left ventricular function, and narrowing of the internal diameter of the supravalvular aortic ridge and atheromatous plaques in the root and ascending aorta may be detected.^56–59^ Of note, echo has lower sensitivity compared with CT scan in detecting calcification of the aortic valve and root. In line with Cuchel et al, we recommend echocardiography with colour and Doppler annually for evaluation of the heart and aorta in all HoFH patients on LA.^1^

We do not recommend IMT for regular monitoring in clinical practice, as there are important limitations: it requires special expertise, the intra-observer variability is high, there are no reported standardized reference values for children, and accepted thresholds for defining the presence and progression of atherosclerosis in children by IMT are lacking.

<u>CT coronary angiography (CTCA) (prospective ECG-triggered):</u> ECG-triggered CTCA is recommended to detect potential hemodynamic obstructive lesions with special focus on the coronary ostia and proximal segments, all risk factors for myocardial infarction. In addition, CTCA enables assessment of both non-calcified and calcified atherosclerotic plaques and thereby identification of subclinical ASCVD. Detection of subclinical ASCVD in HoFH children is an indication for lower LDL-C targets. CTCA is recommended before initiation of LA therapy, to guide treatment decision making and tailor treatment frequency and intensity. With newer therapies, repetitive CTCA may visualize regression of marked plaques in adolescents.^50^ Radiation dose and hence repeated CTCA assessment is acceptable with the latest generation of CT-scanners.^60^ In children, CTCA may result in a median effective dose of only 1 mSv when performed by 128-slice dual source CT.^61^ CTCA on the latest generation of CT-scanners also enables image acquisition with a high temporal resolution that allows for application in new- borns and children without sedation.^61^ If CTCA is not available, one should consider referring patients to the nearest center with such scanner. Follow-up CTCA should be considered if a change in LA therapy intensity is considered. Cuchel et al. recommend ASCVD monitoring by CTCA at least once after the age of 3 years and follow- up CTCA as clinically indicated.^1^ The optimal interval in general is unknown and should be individualized for each patient based on the achievement of LDL-C target levels, presence of ASCVD and results from previous CTCA.

<u>Non-invasive functional tests:</u> cardiac MRI is an accurate modality for assessment of myocardial ischemia and infarction. Cardiac MRI can safely serve as a “gatekeeper” for invasive angiography to avoid negative invasive diagnostic procedures and facilitate procedures for revascularization.^62^ Both SPECT and stress perfusion with PET- CT are alternative imaging modalities used for ischemia detection in adults with chronic coronary syndrome, albeit with associated radiation dose.^63, 64^ Magnetic Resonance coronary angiography is under development, but still has its limitations for clinical use. We recommend to use non-invasive functional tests to detect and evaluate potential coronary ischemia if potentially obstructive atherosclerotic plaque >50% is visible on CTCA.

Cardiac catheterisation and coronary angiography: in severely affected HoFH children with potentially obstructive CAD, cardiac catheterisation and coronary angiography should be performed, for assessment of obstructive CAD and possible invasive interventions [percutaneous coronary intervention (PCI) or coronary artery bypass graft surgery (CABG)].^65^ Invasive test should be guided by a combination of clinical findings and results from anatomical and/or functional testing.

### F. Monitoring side effects

#### Statements

19. We recommend children on LA should be monitored for side effects during each LA session. (ungraded)
20. ACE-inhibitors must not be used during treatment with negatively charged membranes. (level X - strong)

#### Rationale

LA is overall well tolerated and safe in children and adults. Most described side effects of LA in children with HoFH are minor and do not affect the tolerability of the treatment. There are five larger observations including a total of 105 HoFH patients and describing several thousand LA sessions in children.^3, 18, 66–68^ 33-63% of the reported paediatric patients ever experienced an LA associated side effect.^3, 66^ and side effects were described in 0.2 to 2% of the sessions only.^18, 67, 68^ Most frequently reported were hypotension, nausea and technical issues, including problems with vascular access. The need of iron supplementation was reported in one paper in which 3/17 (18%) HoFH children on LA required iron supplementation.^32^ None of the patients discontinued chronic LA treatment due to clinical symptoms or technical difficulties. Patients on LA should be monitored closely, especially during the first LA session, for clinical symptoms such as hypotension, nausea and vomiting. Biochemical follow-up should include next to lipid-metabolism, blood count and iron. In case of unselective plasma exchange, coagulation status and immunoglobulins may be monitored, especially with frequent LA, e.g. twice weekly.

Negatively charged membranes used in LA systems with dextran sulphate columns and modified polyacrylamide gels can induce acute bradykinin release, which in rare cases results in severe anaphylactoid reactions.^21, 69^ These membranes are used in polyacrylate full blood adsorption (DALI) ^70^ and in the dextran sulphate-based systems of Kaneka (LA15 and Liposorber D).^71, 72^ Since plasma kallikrein is activated upon contact with the membranes, ACE-inhibitors are contraindicated in patients receiving LA, especially when systems with negative membranes are used.^69, 70^ Patients must be informed about this contra-indication. If there is no alternative therapy to ACE-inhibitors in single patients, the HELP system for LA may be considered, because this system does not activate the kallikrein-kinin system.^70^ Angiotensin Receptor Blockers (ARB) may be an option to replace ACE-inhibitors.^73, 74^ Also without ACE-inhibitors, patients on these devices can experience reactions that are associated with bradykinin release: nausea and vomiting, flushing, tongue swelling, severe abdominal pain and hypotension.^69^

**Experience of the authors:** to prevent side effects, including bradykinin-release-related side effects, multiple centers prime the membrane with albumin, which seems useful in preventing side effects. In one center, the bradykinin-release related symptoms during LA disappeared when the LA15 membrane was rinsed with saline instead of acetate.

Side effects related to different types of vascular access are described in the vascular access rationale. Side effects related to the anticoagulation used during the LA session are specific for the anticoagulation of choice, heparin or citrate; we therefore refer to the respective literature.

### G. The role of Lipoprotein(a)

#### Statements

21. We suggest measuring Lp(a) levels in all children with homozygous FH at least at the time of diagnosis. (Level C – moderate)

#### Rationale

Besides elevated LDL-C levels, elevated levels of Lipoprotein(a) [Lp(a)] above 50 mg/dL or 125 nmol/L are considered ASCVD risk enhancing.^75–81^ In patients with heterozygous FH, elevated Lp(a) levels are a predictor of ASCVD independent of LDL-C levels. ^82–92^ In children and adults with HoFH, Lp(a) levels are reported to be higher compared to patients with heterozygous FH and normolipidemic controls.^93–96^ Data on whether high Lp(a) is an independent risk factor for those patients is scarce.^13, 16, 97, 98^ Only one out of four available studies reported a significantly increased probability of ASCVD or death in HoFH patients with elevated Lp(a) compared to HoFH patients with non-elevated Lp(a) levels^13^; the other three were negative.^16, 97, 98^

Paediatric guidelines recommend measuring Lp(a) in children with dyslipidaemias including familial hypercholesterolemia.^76, 99–101^ Although the exact impact of Lp(a) as a risk factor for ASCVD in HoFH is unclear, we suggest measuring the Lp(a) levels of children with HoFH at least at the time of diagnosis. Knowing a child’s Lp(a) level can help to define their ASCVD risk beyond LDL-C and improve compliance with a lifetime adoption of a heart-healthy lifestyle.^102^ Secondly, measuring Lp(a) levels provides insight in the true LDL-C levels, because measured LDL-C may be higher than true LDL-C in patients with markedly elevated Lp(a) levels.^103, 104^ Finally, a high Lp(a) may be taken into consideration as an additional indication for initiating LA, in case of LDL-C levels between 3.4 and 7.8 mmol/L (130 and 300 mg/dL). LA is effective in lowering Lp(a) levels (60-70% per session) and its pro-inflammatory oxidized phospholipids in HoFH patients, but its impact on clinical risk reduction of ASCVD is uncertain.^15, 21, 105–114^ In patients with isolated Lp(a), LA-mediated reductions of Lp(a) do seem to reduce the number of events, but these results are mainly derived from observational data and firm conclusions cannot be drawn.^115–122^

Lp(a) levels are predominantly genetically determined: they are relatively low at birth and tend to increase during childhood.^123–125^ We therefore suggest to repeat the measurement of Lp(a) in children with Lp(a) levels close to the ASCVD risk enhancing cut-off at the moment of deciding to start LA.^124^ Since LDL-C levels are determinative for LA management, regular monitoring of Lp(a) will have no impact on the management and, therefore, is not recommended.

### H. The role of Liver transplantation

#### Statement

1. Liver transplantation may be considered in HoFH children with persistently elevated LDL-C levels and progressive ASCVD despite optimal <u>available</u> and tolerated pharmacological treatment and lipoprotein apheresis. (Level C - weak)

#### Rationale

If patients have access to LA and new, highly effective drugs such as PCSK9 inhibitors, lomitapide and evinacumab, LDL-C can be reduced into the target range. However, these treatments may not be widely available. If a patient has persistently elevated LDL-C levels and progressive ASCVD despite optimal LLT treatment including before mentioned new LLT drugs and LA, liver transplantation may be considered as a treatment option.

It is estimated that 75% of the LDL receptors are located in the liver. By replacing the liver with poor LDL receptor function with a normal liver, LDL-C levels decrease to normal levels within a few weeks.^126–128^

The risks of liver transplantation and post-transplant immunosuppressive therapy should be carefully balanced against the risks of persistently elevated LDL-C levels, and the benefits of the new potent LLTs. A complete review of the impact of liver transplantation is beyond the scope of this paper. In short, the risks include surgical complications, in particular cardiovascular hemodynamic instability, hemorrhage, thrombosis of the hepatic artery, hepatic vein, or portal vein and biliary complications; acute and chronic rejection, infection and side effects of immunosuppression.^129, 130^

Data on the effects of liver transplantation on cardiovascular burden are scarce, short-term and usually vaguely described. Regression of coronary artery disease has been described ^127, 131–134^ and survival up to 28 years has been reported.^135^ Besides LDL-C lowering, a yet-to-be proven benefit of liver transplantation is a reduction in Lp(a).^127, 136^ Although liver transplantation reduces the LDL-C levels, some case reports describe progression of aortic stenosis.^127, 128, 137^ In addition to ASCVD benefits, there may be an improvement in the quality of life with liver transplantation compared to weekly or biweekly LA.^138, 139^

#### Research topics to be developed

While developing this paper, we identified gaps in research. Therefore, we suggest the following topics for further research:

i. Specific effect of LA on ASCVD in children with HoFH, especially relative to the novel, potent lipid lowering therapies
ii. Prospective monitoring of different vascular access for LA for efficiency and safety
iii. Cardiac impact of the additional shunt volume of AVF (and AVG)
iv. Impact of LA on quality of life and psychosocial health of children
v. Risk of elevated Lp(a) levels on ASCVD in paediatric HoFH patients

## Data Availability

All data for this manuscript was used from cited articles. No other datasets have been used.

## Acknowledgments

The authors acknowledge the valuable contributions of the members of the voting group who participated in the Delphi technique: Martin P. Bogsrud (Unit for Cardiac and Cardiovascular Genetics, Oslo University Hospital, Oslo, Norway), Eric Bruckert (Pitié-Salpêtrière Hospital and Sorbonne University, Cardio metabolic Institute, Paris, France), Anna Colpo (Therapeutic Apheresis Unit, Department of Transfusion Medicine, Padova University Hospital, Padova, Italy), Marina Cuchel (Division of Translational Medicine and Human Genetics, Department of Medicine, Perelman School of Medicine, University of Pennsylvania, USA), Tomas Freiberger (Centre of Cardiovascular Surgery and Transplantation, Brno, Czech Republic), Jacques Genest (McGill University Health Center, Montreal, Canada), Samual S. Gidding (Geisinger Health, Danvulle, USA), Dieter Haffner (Department of Pediatric Kidney, Liver and Metabolic Diseases, Hannover Medical School, Pediatric Research Center, Hannover, Germany), Ulrich Laufs (Clinic and Policlinic for Cardiology, University Hospital Leipzig, Leipzig, Germany), Christian Loewe (Department Of Bioimaging and Image-Guided Therapy, Medical University of Vienna, Vienna, Austria), Luis Masana (University Rovira & Virgili, Reus-Tarragona, Spain), Brian McCrindle (The Hospital for Sick Children, University of Toronto, Toronto, Canada), Fransesco Sbrana (U.O. Lipoaferesi - Fondazione Toscana “Gabriele Monasterio”, Pisa, Italy), Rukshana Shroff (Great Ormond Street Hospital and Institute of Child Health, University College London, London, United Kingdom), Gilbert Thompson (Imperial College London, London, United Kingdom), Julia Thumfart (Department of Pediatric Gastroenterology, Nephrology and Metabolic Diseases, Charité Universitätsmedizin Berlin, Berlin, Germany), S. Lale Tokgözoğlu (Hacettepe University, Ankara, Turkey), Alpo Vuorio (Department of Medicine, University of Helsinki, Finland), Gerald F. Watts (Medical School, University of Western Australia, and Department of Cardiology, Lipid Disorders Clinic, Royal Perth Hospital, Perth, Australia).

---

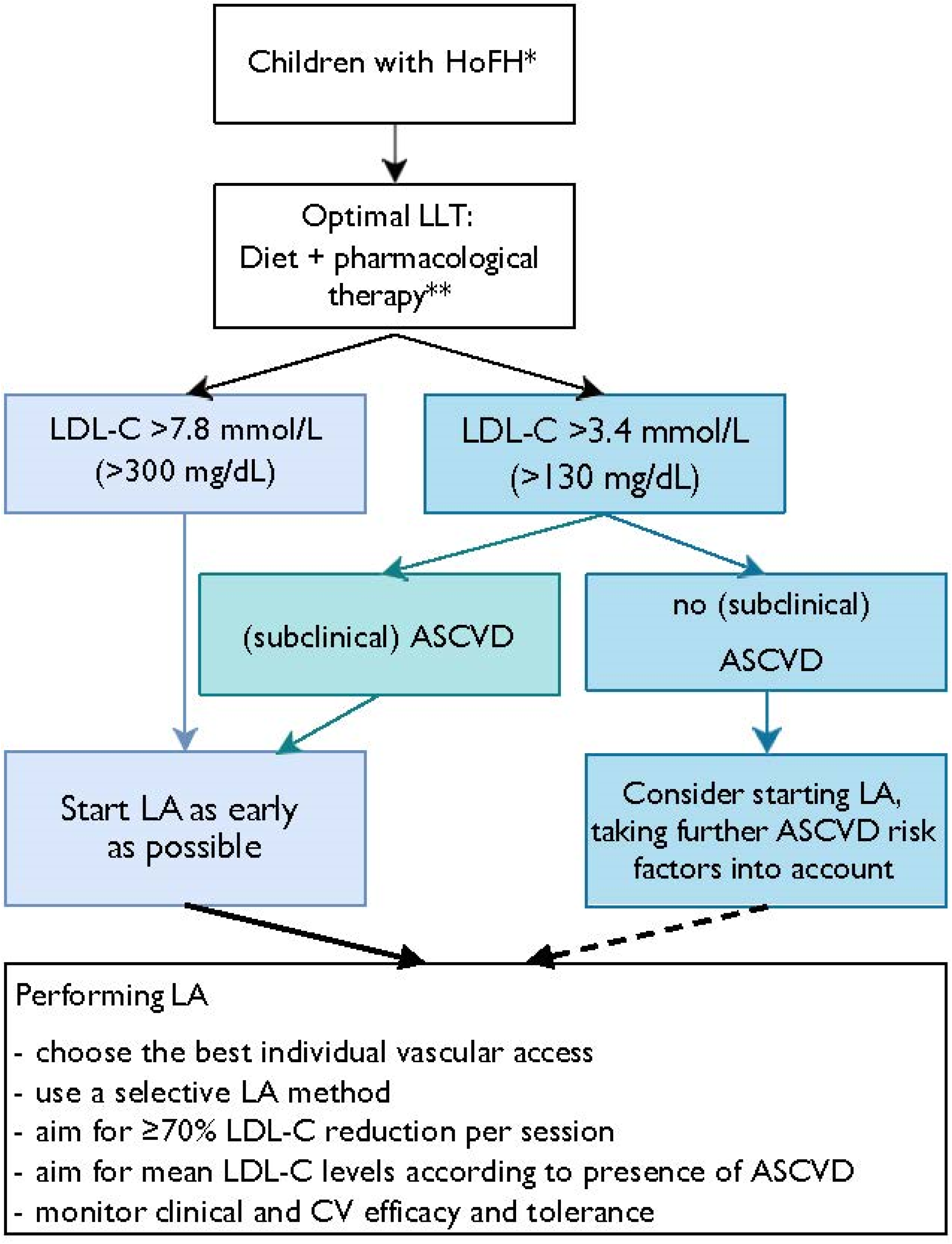

## References

1. Cuchel M, Raal FJ, Hegele RA, Al-Rasadi K, Arca M, Averna M, et al. 2023 Update on European Atherosclerosis Society Consensus Statement on Homozygous Familial Hypercholesterolaemia: new treatments and clinical guidance. Eur Heart J 2023;44(25):2277-2291. doi: 10.1093/eurheartj/ehad197

2. Reijman MD, Kusters DM, Wiegman A. Advances in familial hypercholesterolaemia in children. Lancet Child Adolesc Health 2021;5(9):652–61. doi: 10.1016/s2352-4642(21)00095-x

3. Tromp TR, Hartgers ML, Hovingh GK, Blom DJ, Cuchel M, Raal FJ. Worldwide perspective on homozygous familial hypercholesterolemia diagnosis, treatment and outcome – results from the HICC registry. Atherosclerosis 2021;331:e180–e1. doi: 10.1016/j.atherosclerosis.2021.06.549

4. Luirink IK, Hutten BA, Greber-Platzer S, Kolovou GD, Dann EJ, de Ferranti SD, et al. Practice of lipoprotein apheresis and short-term efficacy in children with homozygous familial hypercholesterolemia: Data from an international registry. Atherosclerosis 2020;299:24–31. doi: 10.1016/j.atherosclerosis.2020.01.031

5. Víšek J, Bláha M, Bláha V, Lášticová M, Lánska M, Andrýs C, et al. Monitoring of up to 15 years effects of lipoprotein apheresis on lipids, biomarkers of inflammation, and soluble endoglin in familial hypercholesterolemia patients. Orphanet J Rare Dis 2021;16(1):110. doi: 10.1186/s13023-021-01749-w

6. Thompson GR. The scientific basis and future of lipoprotein apheresis. Ther Apher Dial 2022;26(1):32–36. doi: 10.1111/1744-9987.13716

7. Gautschi M, Pavlovic M, Nuoffer JM. Fatal myocardial infarction at 4.5 years in a case of homozygous familial hypercholesterolaemia. JIMD Rep 2012;2:45–50. doi: 10.1007/8904_2011_45

8. Guyatt GH, Oxman AD, Kunz R, Atkins D, Brozek J, Vist G, et al. GRADE guidelines: 2. Framing the question and deciding on important outcomes. J Clin Epidemiol 2011;64(4):395–400. doi: 10.1016/j.jclinepi.2010.09.012

9. Improvement SCoQ, Management. Classifying Recommendations for Clinical Practice Guidelines. Pediatrics 2004;114(3):874–7. doi: 10.1542/peds.2004-1260

10. Watts GF, Gidding SS, Hegele RA, Raal FJ, Sturm AC, Jones LK, et al. International Atherosclerosis Society guidance for implementing best practice in the care of familial hypercholesterolaemia. Nat Rev Cardiol 2023;10.1038/s41569-023-00892-0. doi: 10.1038/s41569-023-00892-0

11. 11. Feingold KR, Grunfeld C. Lipoprotein Apheresis. In: Feingold KR, Anawalt B, Boyce A, Chrousos G, de Herder WW, Dhatariya K, et al., editors. Endotext. South Dartmouth (MA): MDText.com, Inc.; February 19, 2023.

12. Daniels SR, Greer FR. Lipid screening and cardiovascular health in childhood. Pediatrics 2008;122(1):198–208. doi: 10.1542/peds.2008-1349

13. Stefanutti C, Pang J, Di Giacomo S, Wu X, Wang X, Morozzi C, et al. A cross-national investigation of cardiovascular survival in homozygous familial hypercholesterolemia: The Sino-Roman Study. J Clin Lipidol 2019;13(4):608–17. doi: 10.1016/j.jacl.2019.05.002

14. Lischka J, Arbeiter K, de Gier C, Willfort-Ehringer A, Walleczek N-K, Gellai R, et al. Vascular access for lipid apheresis: a challenge in young children with homozygous familial hypercholesterolemia. BMC Pediatrics 2022;22(1):131. doi: 10.1186/s12887-022-03192-7

15. Stefanutti C, Di Giacomo S, Vivenzio A, Colloridi V, Bosco G, Berni A, et al. Low-density lipoprotein apheresis in a patient aged 3.5 years. Acta Paediatr 2001;90(6):694–701. doi: 10.1080/080352501750258793

16. Taylan C, Driemeyer J, Schmitt CP, Pape L, Buscher R, Galiano M, et al. Cardiovascular Outcome of Pediatric Patients With Bi-Allelic (Homozygous) Familial Hypercholesterolemia Before and After Initiation of Multimodal Lipid Lowering Therapy Including Lipoprotein Apheresis. Am J Cardiol 2020;136:38–48. doi: 10.1016/j.amjcard.2020.09.015

17. Fernández-Fuertes LF, Tapia Martín M, Nieves Plá I, Novoa Mogollón FJ, Díaz Cremades J. Low-Density Lipoprotein Apheresis Using Double Filtration Plasmapheresis: 27-Month Use in a Child With Homozygous Familial Hypercholesterolemia. Therapeutic Apheresis and Dialysis 2010;14(5):484–5. doi: 10.1111/j.1744-9987.2010.00839.x

18. Coker M, Ucar SK, Simsek DG, Darcan S, Bak M, Can S. Low density lipoprotein apheresis in pediatric patients with homozygous familial hypercholesterolemia. Ther Apher Dial 2009;13(2):121–8. doi: 10.1111/j.1744-9987.2009.00666.x

19. Lefort B, Saheb S, Bruckert E, Giraud C, Hequet O, Hankard R. Impact of LDL apheresis on aortic root atheroma in children with homozygous familial hypercholesterolemia. Atherosclerosis 2015;239(1):158–62. doi: 10.1016/j.atherosclerosis.2015.01.007

20. Luirink IK, Determeijer J, Hutten BA, Wiegman A, Bruckert E, Schmitt CP, et al. Efficacy and safety of lipoprotein apheresis in children with homozygous familial hypercholesterolemia: A systematic review. J Clin Lipidol 2019;13(1):31–9. doi: 10.1016/j.jacl.2018.10.011

21. Hudgins LC, Kleinman B, Scheuer A, White S, Gordon BR. Long-term safety and efficacy of low-density lipoprotein apheresis in childhood for homozygous familial hypercholesterolemia. Am J Cardiol 2008;102(9):1199–204. doi: 10.1016/j.amjcard.2008.06.049

22. Buonuomo PS, Macchiaiolo M, Leone G, Valente P, Mastrogiorgio G, Gnazzo M, et al. Treatment of homozygous familial hypercholesterolaemia in paediatric patients: A monocentric experience. Eur J Prev Cardiol 2018;25(10):1098–105. doi: doi:10.1177/2047487318776836

23. Padmanabhan A, Connelly-Smith L, Aqui N, Balogun RA, Klingel R, Meyer E, et al. Guidelines on the Use of Therapeutic Apheresis in Clinical Practice - Evidence-Based Approach from the Writing Committee of the American Society for Apheresis: The Eighth Special Issue. J Clin Apher 2019;34(3):171–354. doi: 10.1002/jca.21705

24. Bambauer R, Bambauer C, Lehmann B, Latza R, Schiel R. LDL-Apheresis: Technical and Clinical Aspects. Scientific World Journal 2012;2012:314283. doi: 10.1100/2012/314283

25. Klingel R, Fassbender T, Fassbender C, Göhlen B. From Membrane Differential Filtration to Lipidfiltration: Technological Progress in Low-density Lipoprotein Apheresis. Ther Apher Dial 2003;7(3):350–8. doi: 10.1046/j.1526-0968.2003.00062.x

26. Drouin-Chartier JP, Tremblay AJ, Bergeron J, Pelletier M, Laflamme N, Lamarche B, et al. Comparison of two low-density lipoprotein apheresis systems in patients with homozygous familial hypercholesterolemia. J Clin Apher 2016;31(4):359–67. doi: 10.1002/jca.21406

27. Kopprasch S, Bornstein SR, Bergmann S, Graessler J, Julius U. Long-term therapeutic efficacy of lipoprotein apheresis on circulating oxidative stress parameters--A comparison of two different apheresis techniques. Atheroscler Suppl 2015;18:80–4. doi: 10.1016/j.atherosclerosissup.2015.02.016

28. Roeseler E, Julius U, Heigl F, Spitthoever R, Heutling D, Breitenberger P, et al. Lipoprotein Apheresis for Lipoprotein(a)-Associated Cardiovascular Disease. Arterioscler Thromb Vasc Biol 2016;36(9):2019–27. doi: doi: 10.1161/ATVBAHA.116.307983

29. Gokay S, Kendirci M, Kaynar L, Solmaz M, Cetin A, Kardas F, et al. Long-term efficacy of lipoprotein apheresis in the management of familial hypercholesterolaemia: Application of two different apheresis techniques in childhood. Transfus Apher Sci 2016;54(2):282–8. doi: 10.1016/j.transci.2015.10.015

30. Albayrak M, Yıldız A, Ateş N, Pala Ç. The efficacy of double filtration plasmapheresis in the treatment of homozygous familial hypercholesterolemia: A single-center experience. Transfus Apher Sci 2019;58(1):61–4. doi: 10.1016/j.transci.2018.11.007

31. Shin HS, Towbin AJ, Zhang B, Johnson ND, Goldstein SL. Venous thrombosis and stenosis after peripherally inserted central catheter placement in children. Pediatr Radiol 2017;47(12):1670–5. doi: 10.1007/s00247-017-3915-9

32. Klaus G, Taylan C, Büscher R, Schmitt CP, Pape L, Oh J, et al. Multimodal lipid-lowering treatment in pediatric patients with homozygous familial hypercholesterolemia—target attainment requires further increase of intensity. Pediatr Nephrol 2018;33(7):1199–208. doi: doi:10.1007/s00467-018-3906-6

33. Dann EJ, Shamir R, Mashiach T, Shaoul R, Badian A, Stravets T, et al. Early-onset plasmapheresis and LDL- apheresis provide better disease control for pediatric homozygous familial hypercholesterolemia than HMG-CoA reductase inhibitors and ameliorate atherosclerosis. Transfus Apher Sci 2013;49(2):268–77. doi: 10.1016/j.transci.2013.05.001

34. Borzych-Duzalka D, Shroff R, Ariceta G, Yap Y-C, Paglialonga F, Xu H, et al. Vascular Access Choice, Complications, and Outcomes in Children on Maintenance Hemodialysis: Findings From the International Pediatric Hemodialysis Network (IPHN) Registry. Am J Kidney Dis 2019;74(2):193–202. doi: 10.1053/j.ajkd.2019.02.014

35. Rao NN, Stokes MB, Rajwani A, Ullah S, Williams K, King D, et al. Effects of Arteriovenous Fistula Ligation on Cardiac Structure and Function in Kidney Transplant Recipients. Circulation 2019;139(25):2809–18. doi: 10.1161/CIRCULATIONAHA.118.038505

36. Cridlig J, Selton-Suty C, Alla F, Chodek A, Pruna A, Kessler M, et al. Cardiac impact of the arteriovenous fistula after kidney transplantation: a case-controlled, match-paired study. Transpl Int 2008;21(10):948–54. doi: 10.1111/j.1432-2277.2008.00707.x

37. Çakıcı EK, Çakıcı M, Gümüş F, Tan Kürklü TS, Yazılıtaş F, Örün UA, et al. Effects of hemodialysis access type on right heart geometry in adolescents. J Vasc Access 2020;21(5):658–64. doi: 10.1177/1129729819897454

38. Alkhouli M, Sandhu P, Boobes K, Hatahet K, Raza F, Boobes Y. Cardiac complications of arteriovenous fistulas in patients with end-stage renal disease. Nefrología 2015;35(3):234–45. doi: 10.1016/j.nefro.2015.03.001

39. Malik J. Heart disease in chronic kidney disease - review of the mechanisms and the role of dialysis access. J Vasc Access 2018;19(1):3–11. doi: 10.5301/jva.5000815

40. Chand DH, Bednarz D, Eagleton M, Krajewski L. A vascular access team can increase AV fistula creation in pediatric ESRD patients: a single center experience. Semin Dial 2009;22(6):679–83. doi: 10.1111/j.1525-139X.2009.00638.x

41. Shroff R, Calder F, Bakkaloğlu S, Nagler EV, Stuart S, Stronach L, et al. Vascular access in children requiring maintenance haemodialysis: a consensus document by the European Society for Paediatric Nephrology Dialysis Working Group. Nephrol Dial Transplant 2019;34(10):1746–65. doi: 10.1093/ndt/gfz011

42. Thom KE, Hölzenbein T, Jones N, Zwiauer K, Streif W, Gattringer S, et al. Arteriovenous shunts as venous access in children with haemophilia. Haemophilia 2018;24(3):429–35. doi: 10.1111/hae.13433

43. Karava V, Jehanno P, Kwon T, Deschênes G, Macher MA, Bourquelot P. Autologous arteriovenous fistulas for hemodialysis using microsurgery techniques in children weighing less than 20 kg. Pediatr Nephrol 2018;33(5):855–62. doi: 10.1007/s00467-017-3854-6

44. Katzmann JL, Gouni-Berthold I, Laufs U. PCSK9 Inhibition: Insights From Clinical Trials and Future Prospects. Front Physiol 2020;11:595819. doi: 10.3389/fphys.2020.595819

45. Ference BA, Ginsberg HN, Graham I, Ray KK, Packard CJ, Bruckert E, et al. Low-density lipoproteins cause atherosclerotic cardiovascular disease. 1. Evidence from genetic, epidemiologic, and clinical studies. A consensus statement from the European Atherosclerosis Society Consensus Panel. Eur Heart J 2017;38(32):2459–72. doi: 10.1093/eurheartj/ehx144

46. Cuchel M, Bruckert E, Ginsberg HN, Raal FJ, Santos RD, Hegele RA, et al. Homozygous familial hypercholesterolaemia: new insights and guidance for clinicians to improve detection and clinical management. A position paper from the Consensus Panel on Familial Hypercholesterolaemia of the European Atherosclerosis Society. Eur Heart J 2014;35(32):2146–57. doi: 10.1093/eurheartj/ehu274

47. Santos RD, Ruzza A, Hovingh GK, Wiegman A, Mach F, Kurtz CE, et al. Evolocumab in Pediatric Heterozygous Familial Hypercholesterolemia. N Engl J Med 2020;383(14):1317–1327. doi: 10.1056/NEJMoa2019910

48. Buonuomo PS, Mastrogiorgio G, Leone G, Rana I, Gonfiantini MV, Macchiaiolo M, et al. Evolocumab in the management of children &lt;10 years of age affected by homozygous familial hypercholesterolemia. Atherosclerosis 2021;324:148–50. doi: doi:10.1016/j.atherosclerosis.2021.03.026

49. Raal FJ, Rosenson RS, Reeskamp LF, Hovingh GK, Kastelein JJP, Rubba P, et al. Evinacumab for Homozygous Familial Hypercholesterolemia. N Engl J Med 2020;383(8):711–20. doi: 10.1056/NEJMoa2004215

50. Reeskamp LF, Nurmohamed NS, Bom MJ, Planken RN, Driessen RS, van Diemen PA, et al. Marked plaque regression in homozygous familial hypercholesterolemia. Atherosclerosis 2021;327:13–7. doi: 10.1016/j.atherosclerosis.2021.04.014

51. Kroon AA, van’t Hof MA, Demacker PN, Stalenhoef AF. The rebound of lipoproteins after LDL-apheresis. Kinetics and estimation of mean lipoprotein levels. Atherosclerosis 2000;152(2):519–26. doi: 10.1016/s0021-9150(00)00371-3

52. Thompson GR, Barbir M, Davies D, Dobral P, Gesinde M, Livingston M, et al. Efficacy criteria and cholesterol targets for LDL apheresis. Atherosclerosis 2010;208(2):317–21. doi: 10.1016/j.atherosclerosis.2009.06.010

53. Ring L, Shah BN, Bhattacharyya S, Harkness A, Belham M, Oxborough D, et al. Echocardiographic assessment of aortic stenosis: a practical guideline from the British Society of Echocardiography. Echo Res Pract 2021;8(1):G19–g59. doi: 10.1530/erp-20-0035

54. Belanger AM, Akioyamen LE, Ruel I, Hales L, Genest J. Aortic stenosis in homozygous familial hypercholesterolaemia: a paradigm shift over a century. Eur Heart J 2022;43(34):3227–39. doi: 10.1093/eurheartj/ehac339

55. Kawaguchi A, Yutani C, Yamamoto A. Hypercholesterolemic valvulopathy: an aspect of malignant atherosclerosis. Ther Apher Dial 2003;7(4):439–43. doi: 10.1046/j.1526-0968.2003.00075.x

56. Awan Z, Alrasadi K, Francis GA, Hegele RA, McPherson R, Frohlich J, et al. Vascular calcifications in homozygote familial hypercholesterolemia. Arterioscler Thromb Vasc Biol 2008;28(4):777–85. doi: 10.1161/ATVBAHA.107.160408

57. Hoeg JM, Feuerstein IM, Tucker EE. Detection and quantitation of calcific atherosclerosis by ultrafast computed tomography in children and young adults with homozygous familial hypercholesterolemia. Arterioscler Thromb 1994;14(7):1066–74. doi: 10.1161/01.atv.14.7.1066

58. Labombarda F, Castelnuovo S, Goularas D, Sirtori CR. Status and potential clinical value of a transthoracic evaluation of the coronary arteries. Cardiovasc Ultrasound 2016;14:5. doi: 10.1186/s12947-016-0048-5

59. Kawaguchi A, Miyatake K, Yutani C, Beppu S, Tsushima M, Yamamura T, et al. Characteristic cardiovascular manifestation in homozygous and heterozygous familial hypercholesterolemia. Am Heart J 1999;137(3):410–8. doi: 10.1016/s0002-8703(99)70485-0

60. Sabarudin A, Sun Z, Ng KH. Coronary computed tomography angiography with prospective electrocardiography triggering: a systematic review of image quality and radiation dose. Singapore Med J 2013;54(1):15–23. doi: 10.11622/smedj.2013005

61. Ghoshhajra BB, Lee AM, Engel L-C, Celeng C, Kalra MK, Brady TJ, et al. Radiation Dose Reduction in Pediatric Cardiac Computed Tomography: Experience from a Tertiary Medical Center. Pediatr Cardiol 2014;35(1):171–9. doi: 10.1007/s00246-013-0758-5

62. Ge Y, Deva DP, Connelly KA, Yan AT. Stress cardiac MRI in stable coronary artery disease. Curr Opin Cardiol 2020;35(5):566–73. doi: 10.1097/HCO.0000000000000776

63. Berman DS, Shaw LJ, Min JK, Hachamovitch R, Abidov A, Germano G, et al. SPECT/PET myocardial perfusion imaging versus coronary CT angiography in patients with known or suspected CAD. Q J Nucl Med Mol Imaging 2010;54(2):177–200.

64. Knuuti J, Wijns W, Saraste A, Capodanno D, Barbato E, Funck-Brentano C, et al. 2019 ESC Guidelines for the diagnosis and management of chronic coronary syndromes. Eur Heart J 2020;41(3):407-77. doi: 10.1093/eurheartj/ehz425

65. Revaiah PC, Bootla D, Vemuri KS, Nevali KP, Ghosh S, Sharma YP, et al. Left main revascularization with optical coherence tomography in a young male with newly diagnosed homozygous familial hypercholesterolemia. J Cardiol Cases 2022;25(1):14–8. doi: 10.1016/j.jccase.2021.05.011

66. Palcoux JB, Atassi-Dumont M, Lefevre P, Hequet O, Schlienger JL, Brignon P, et al. Low-density lipoprotein apheresis in children with familial hypercholesterolemia: follow-up to 21 years. Ther Apher Dial 2008;12(3):195–201. doi: 10.1111/j.1744-9987.2008.00574.x

67. Bulut M, Nisli K, DIndar A. The effect of DALI lipid apheresis in the prognosis of homozygous familial hypercholesterolemia: Seven patients’ experience at a DALI apheresis center. Ann Pediatr Cardiol 2020;13(2):111–6. doi:10.4103/apc.APC_56_19

68. Stefanutti C, Vivenzio A, Di Giacomo S, Mazzarella B, Bosco G, Berni A. Aorta and coronary angiographic follow-up of children with severe hypercholesterolemia treated with low-density lipoprotein apheresis. Transfusion 2009;49(7):1461–70. doi: 10.1111/j.1537-2995.2009.02135.x

69. Alexander E, Moriarty PM, Wilk B, Eliaz I. Establishing low-density lipoprotein apheresis tolerability in patients with prior anaphylactoid reactions to lipoprotein apheresis using magnesium sulfate. J Clin Apher 2021;36(3):437–42. doi: 10.1002/jca.21884

70. Krieter DH, Steinke J, Kerkhoff M, Fink E, Lemke HD, Zingler C, et al. Contact activation in low-density lipoprotein apheresis systems. Artif Organs 2005;29(1):47–52. doi: 10.1111/j.1525-1594.2004.29007.x

71. Keller C, Grützmacher P, Bahr F, Schwarzbeck A, Kroon AA, Kiral A. LDL-apheresis with dextran sulphate and anaphylactoid reactions to ACE inhibitors. Lancet 1993;341(8836):60-1. doi: 10.1016/0140-6736(93)92542-2

72. Kroon AA, Mol MJTM, Stalenhoef AFH. ACE inhibitors and LDL-apheresis with dextran sulphate adsorption. The Lancet 1992;340(8833):1476. doi: 10.1016/0140-6736(92)92673-4

73. Sinzinger H, Chehne F, Ferlitsch A, Oguogho A. Angiotensin receptor antagonists during dextran sulfate LDL-apheresis are safe. Thromb Res 2000;100(1):43–6. doi: 10.1016/s0049-3848(00)00301-7

74. Bosch T, Wendler T. Efficacy and safety of DALI-LDL-apheresis in two patients treated with the angiotensin II-receptor 1 antagonist losartan. Ther Apher Dial 2004;8(4):269–74. doi: 10.1111/j.1526-0968.2004.00162.x

75. Grundy SM, Stone NJ, Bailey AL, Beam C, Birtcher KK, Blumenthal RS, et al. 2018 AHA/ACC/AACVPR/AAPA/ABC/ACPM/ADA/AGS/APhA/ASPC/NLA/PCNA Guideline on the Management of Blood Cholesterol: A Report of the American College of Cardiology/American Heart Association Task Force on Clinical Practice Guidelines. J Am Coll Cardiol 2019;73(24):e285-e350. doi: 10.1016/j.jacc.2018.11.003

76. Wilson DP, Jacobson TA, Jones PH, Koschinsky ML, McNeal CJ, Nordestgaard BG, et al. Use of Lipoprotein(a) in clinical practice: A biomarker whose time has come. A scientific statement from the National Lipid Association. J Clin Lipidol 2019;13(3):374–92. doi: 10.1016/j.jacl.2019.04.010

77. Madsen CM, Kamstrup PR, Langsted A, Varbo A, Nordestgaard BG. Lipoprotein(a)-Lowering by 50 mg/dL (105 nmol/L) May Be Needed to Reduce Cardiovascular Disease 20% in Secondary Prevention: A Population-Based Study. Arterioscler Thromb Vasc Biol 2020;40(1):255–66. doi: 10.1161/atvbaha.119.312951

78. Erqou S, Kaptoge S, Perry PL, Di Angelantonio E, Thompson A, White IR, et al. Lipoprotein(a) concentration and the risk of coronary heart disease, stroke, and nonvascular mortality. Jama 2009;302(4):412–23. doi: 10.1001/jama.2009.1063

79. Littmann K, Hagström E, Häbel H, Bottai M, Eriksson M, Parini P, et al. Plasma lipoprotein(a) measured in the routine clinical care is associated to atherosclerotic cardiovascular disease during a 14-year follow-up. Eur J Prev Cardiol 2022;28(18):2038–47. doi: 10.1093/eurjpc/zwab016

80. Patel AP, Wang M, Pirruccello JP, Ellinor PT, Ng K, Kathiresan S, et al. Lp(a) (Lipoprotein[a]) Concentrations and Incident Atherosclerotic Cardiovascular Disease: New Insights From a Large National Biobank. Arterioscler Thromb Vasc Biol 2021;41(1):465–74. doi: 10.1161/atvbaha.120.315291

81. Reyes-Soffer G, Ginsberg HN, Berglund L, Duell PB, Heffron SP, Kamstrup PR, et al. Lipoprotein(a): A Genetically Determined, Causal, and Prevalent Risk Factor for Atherosclerotic Cardiovascular Disease: A Scientific Statement From the American Heart Association. Arterioscler Thromb Vasc Biol 2022;42(1):e48–e60. doi: 10.1161/atv.0000000000000147

82. Allard MD, Saeedi R, Yousefi M, Frohlich J. Risk stratification of patients with familial hypercholesterolemia in a multi-ethnic cohort. Lipids Health Dis 2014;13:65. doi: 10.1186/1476-511x-13-65

83. Alonso R, Andres E, Mata N, Fuentes-Jiménez F, Badimón L, López-Miranda J, et al. Lipoprotein(a) levels in familial hypercholesterolemia: an important predictor of cardiovascular disease independent of the type of LDL receptor mutation. J Am Coll Cardiol 2014;63(19):1982–9. doi: 10.1016/j.jacc.2014.01.063

84. Chan DC, Pang J, Hooper AJ, Burnett JR, Bell DA, Bates TR, et al. Elevated lipoprotein(a), hypertension and renal insufficiency as predictors of coronary artery disease in patients with genetically confirmed heterozygous familial hypercholesterolemia. Int J Cardiol 2015;201:633–8. doi: 10.1016/j.ijcard.2015.08.146

85. Ellis KL, Pang J, Chieng D, Bell DA, Burnett JR, Schultz CJ, et al. Elevated lipoprotein(a) and familial hypercholesterolemia in the coronary care unit: Between Scylla and Charybdis. Clin Cardiol 2018;41(3):378–84. doi: 10.1002/clc.22880

86. Holmes DT, Schick BA, Humphries KH, Frohlich J. Lipoprotein(a) is an independent risk factor for cardiovascular disease in heterozygous familial hypercholesterolemia. Clin Chem 2005;51(11):2067–73. doi: 10.1373/clinchem.2005.055228

87. Jansen AC, van Aalst-Cohen ES, Tanck MW, Trip MD, Lansberg PJ, Liem AH, et al. The contribution of classical risk factors to cardiovascular disease in familial hypercholesterolaemia: data in 2400 patients. J Intern Med 2004;256(6):482–90. doi: 10.1111/j.1365-2796.2004.01405.x

88. Langsted A, Kamstrup PR, Benn M, Tybjærg-Hansen A, Nordestgaard BG. High lipoprotein(a) as a possible cause of clinical familial hypercholesterolaemia: a prospective cohort study. Lancet Diabetes Endocrinol 2016;4(7):577–87. doi: 10.1016/s2213-8587(16)30042-0

89. Li S, Wu NQ, Zhu CG, Zhang Y, Guo YL, Gao Y, et al. Significance of lipoprotein(a) levels in familial hypercholesterolemia and coronary artery disease. Atherosclerosis 2017;260:67–74. doi: 10.1016/j.atherosclerosis.2017.03.021

90. Nenseter MS, Lindvig HW, Ueland T, Langslet G, Ose L, Holven KB, et al. Lipoprotein(a) levels in coronary heart disease-susceptible and -resistant patients with familial hypercholesterolemia. Atherosclerosis 2011;216(2):426–32. doi: 10.1016/j.atherosclerosis.2011.02.007

91. Pavanello C, Pirazzi C, Bjorkman K, Sandstedt J, Tarlarini C, Mosca L, et al. Individuals with familial hypercholesterolemia and cardiovascular events have higher circulating Lp(a) levels. J Clin Lipidol 2019;13(5):778–87.e6. doi: 10.1016/j.jacl.2019.06.011

92. de Boer LM, Wiegman A, Kroon J, Tsimikas S, Yeang C, Peletier MC, et al. Lipoprotein(a) and carotid intima-media thickness in children with familial hypercholesterolaemia in the Netherlands: a 20-year follow-up study. Lancet Diabetes Endocrinol 2023 ;11(9):667–674. doi: 10.1016/S2213-8587(23)00156-0

93. Guo HC, Chapman MJ, Bruckert E, Farriaux JP, De Gennes JL. Lipoprotein Lp(a) in homozygous familial hypercholesterolemia: density profile, particle heterogeneity and apolipoprotein(a) phenotype. Atherosclerosis 1991;86(1):69–83. doi: 10.1016/0021-9150(91)90100-h

94. Kraft HG, Lingenhel A, Raal FJ, Hohenegger M, Utermann G. Lipoprotein(a) in homozygous familial hypercholesterolemia. Arterioscler Thromb Vasc Biol 2000;20(2):522–8. doi: 10.1161/01.atv.20.2.522

95. Sjouke B, Yahya R, Tanck MWT, Defesche JC, de Graaf J, Wiegman A, et al. Plasma lipoprotein(a) levels in patients with homozygous autosomal dominant hypercholesterolemia. J Clin Lipidol 2017;11(2):507–14. doi: 10.1016/j.jacl.2017.02.010

96. de Boer LM, Reijman MD, Hutten BA, Wiegman A. Lipoprotein(a) levels in children with homozygous familial hypercholesterolaemia: A cross-sectional study. J Clin Lipidol 2023;17(3):415–419. doi: 10.1016/j.jacl.2023.03.010

97. Bruckert E, Kalmykova O, Bittar R, Carreau V, Béliard S, Saheb S, et al. Long-term outcome in 53 patients with homozygous familial hypercholesterolaemia in a single centre in France. Atherosclerosis 2017;257:130–7. doi: 10.1016/j.atherosclerosis.2017.01.015

98. Thompson GR, Blom DJ, Marais AD, Seed M, Pilcher GJ, Raal FJ. Survival in homozygous familial hypercholesterolaemia is determined by the on-treatment level of serum cholesterol. European Heart Journal 2018;39(14):1162–8. doi: 10.1093/eurheartj/ehx317

99. Grundy SM, Stone NJ, Bailey AL, Beam C, Birtcher KK, Blumenthal RS, et al. 2018 AHA/ACC/AACVPR/AAPA/ABC/ACPM/ADA/AGS/APhA/ASPC/NLA/PCNA Guideline on the Management of Blood Cholesterol: A Report of the American College of Cardiology/American Heart Association Task Force on Clinical Practice Guidelines. Circulation 2019;139(25):e1082-e143. doi: 10.1161/cir.0000000000000625

100. Bjelakovic B, Stefanutti C, Reiner Ž, Watts GF, Moriarty P, Marais D, et al. Risk Assessment and Clinical Management of Children and Adolescents with Heterozygous Familial Hypercholesterolaemia. A Position Paper of the Associations of Preventive Pediatrics of Serbia, Mighty Medic and International Lipid Expert Panel. J Clin Med 2021;10(21):4930. doi: 10.3390/jcm10214930

101. De Jesus JM. Expert Panel on Integrated Guidelines for Cardiovascular Health and Risk Reduction in Children and Adolescents: Summary Report. Pediatrics 2011;128(Supplement_5):S213-S56. doi: 10.1542/peds.2009-2107C

102. Perrot N, Verbeek R, Sandhu M, Boekholdt SM, Hovingh GK, Wareham NJ, et al. Ideal cardiovascular health influences cardiovascular disease risk associated with high lipoprotein(a) levels and genotype: The EPIC-Norfolk prospective population study. Atherosclerosis 2017;256:47–52. doi: 10.1016/j.atherosclerosis.2016.11.010

103. Kinpara K, Okada H, Yoneyama A, Okubo M, Murase T. Lipoprotein(a)-cholesterol: a significant component of serum cholesterol. Clin Chim Acta 2011;412(19-20):1783–7. doi: 10.1016/j.cca.2011.05.036

104. Yeang C, Witztum JL, Tsimikas S. Novel method for quantification of lipoprotein(a)-cholesterol: implications for improving accuracy of LDL-C measurements. J Lipid Res 2021;62:100053. doi: 10.1016/j.jlr.2021.100053

105. Stefanutti C, Vivenzio A, Colombo C, Di Giacomo S, Mazzarella B, Berni A, et al. Treatment of homozygous and double heterozygous familial hypercholesterolemic children with LDL-apheresis. Int J Artif Organs 1995;18(2):103–10.

106. Taylan C, Schlune A, Meissner T, Ažukaitis K, Udink Ten Cate FE, Weber LT. Disease control via intensified lipoprotein apheresis in three siblings with familial hypercholesterolemia. J Clin Lipidol 2016;10(6):1303–10. doi: 10.1016/j.jacl.2016.08.006

107. Zwiener RJ, Uauy R, Petruska ML, Huet BA. Low-density lipoprotein apheresis as long-term treatment for children with homozygous familial hypercholesterolemia. J Pediatr 1995;126(5 Pt 1):728-35. doi: 10.1016/s0022-3476(95)70400-0

108. Arai K, Orsoni A, Mallat Z, Tedgui A, Witztum JL, Bruckert E, et al. Acute impact of apheresis on oxidized phospholipids in patients with familial hypercholesterolemia. J Lipid Res 2012;53(8):1670–8. doi: 10.1194/jlr.P027235

109. Græsdal A, Bogsrud MP, Holven KB, Nenseter MS, Narverud I, Langslet G, et al. Apheresis in homozygous familial hypercholesterolemia: the results of a follow-up of all Norwegian patients with homozygous familial hypercholesterolemia. J Clin Lipidol 2012;6(4):331–9. doi: 10.1016/j.jacl.2012.03.004

110. Lasunción MA, Teruel JL, Alvarez JJ, Carrero P, Ortuño J, Gómez-Coronado D. Changes in lipoprotein(a), LDL-cholesterol and apolipoprotein B in homozygous familial hypercholesterolaemic patients treated with dextran sulfate LDL-apheresis. Eur J Clin Invest 1993;23(12):819–26. doi: 10.1111/j.1365-2362.1993.tb00736.x

111. Stefanutti C, Di Giacomo S, Vivenzio A, Isacchi GC, Masella R, Caprari P, et al. Acute and long-term effects of low-density lipoprotein (LDL)-apheresis on oxidative damage to LDL and reducing capacity of erythrocytes in patients with severe familial hypercholesterolaemia. Clin Sci (Lond) 2001;100(2):191–8.

112. Krebs A, Krebs K, Keller F. Retrospective Comparison of 5 different Methods for Long-Term LDL-Apheresis in 20 Patients between 1986 and 2001. Int J Artif Organs 2004;27(2):137–48. doi: 10.1177/039139880402700209

113. Waldmann E, Parhofer KG. Lipoprotein apheresis to treat elevated lipoprotein (a). J Lipid Res 2016;57(10):1751–7. doi: 10.1194/jlr.R056549

114. Greco MF, Sirtori CR, Corsini A, Ezhov M, Sampietro T, Ruscica M. Lipoprotein(a) Lowering-From Lipoprotein Apheresis to Antisense Oligonucleotide Approach. J Clin Med 2020;9(7):2103. doi: 10.3390/jcm9072103

115. Groß E, Hohenstein B, Julius U. Effects of Lipoprotein apheresis on the Lipoprotein(a) levels in the long run. Atheroscler Suppl 2015;18:226–32. doi: 10.1016/j.atherosclerosissup.2015.02.033

116. Jaeger BR, Richter Y, Nagel D, Heigl F, Vogt A, Roeseler E, et al. Longitudinal cohort study on the effectiveness of lipid apheresis treatment to reduce high lipoprotein(a) levels and prevent major adverse coronary events. Nat Clin Pract Cardiovasc Med 2009;6(3):229–39. doi: 10.1038/ncpcardio1456

117. Moriarty PM, Gray JV, Gorby LK. Lipoprotein apheresis for lipoprotein(a) and cardiovascular disease. J Clin Lipidol 2019;13(6):894–900. doi: 10.1016/j.jacl.2019.09.010

118. Pokrovsky SN, Afanasieva OI, Safarova MS, Balakhonova TV, Matchin YG, Adamova IYU, et al. Specific Lp(a) apheresis: A tool to prove lipoprotein(a) atherogenicity. Atheroscler Suppl 2017;30:166–73. doi: 10.1016/j.atherosclerosissup.2017.05.004

119. Roeseler E, Julius U, Heigl F, Spitthoever R, Heutling D, Breitenberger P, et al. Lipoprotein Apheresis for Lipoprotein(a)-Associated Cardiovascular Disease: Prospective 5 Years of Follow-Up and Apolipoprotein(a) Characterization. Arterioscler Thromb Vasc Biol 2016;36(9):2019–27. doi: 10.1161/atvbaha.116.307983

120. Rosada A, Kassner U, Vogt A, Willhauck M, Parhofer K, Steinhagen-Thiessen E. Does regular lipid apheresis in patients with isolated elevated lipoprotein(a) levels reduce the incidence of cardiovascular events? Artif Organs 2014;38(2):135–41. doi: 10.1111/aor.12135

121. von Dryander M, Fischer S, Passauer J, Müller G, Bornstein SR, Julius U. Differences in the atherogenic risk of patients treated by lipoprotein apheresis according to their lipid pattern. Atheroscler Suppl 2013;14(1):39–44. doi: 10.1016/j.atherosclerosissup.2012.10.005

122. Stefanutti C, Di Giacomo S, Mazzarella B, Castelli A. LDL apheresis: a novel technique (LIPOCOLLECT 200). Artif Organs 2009;33(12):1103–8. doi: 10.1111/j.1525-1594.2009.00959.x

123. Routi T, Rönnemaa T, Viikari JS, Leino A, Välimäki IA, Simell OG. Tracking of serum lipoprotein (a) concentration and its contribution to serum cholesterol values in children from 7 to 36 months of age in the STRIP Baby Study. Special Turku Coronary Risk Factor Intervention Project for Babies. Ann Med 1997;29(6):541–7. doi: 10.3109/07853899709007479

124. de Boer LM, Hof MH, Wiegman A, Stroobants AK, Kastelein JJP, Hutten BA. Lipoprotein(a) levels from childhood to adulthood: Data in nearly 3,000 children who visited a pediatric lipid clinic. Atherosclerosis 2022;349:227–232. doi: 10.1016/j.atherosclerosis.2022.03.004

125. Strandkjær N, Hansen MK, Nielsen ST, Frikke-Schmidt R, Tybjærg-Hansen A, Nordestgaard BG, et al. Lipoprotein(a) Levels at Birth and in Early Childhood: The COMPARE Study. J Clin Endocrinol Metab 2022;107(2):324–35. doi: 10.1210/clinem/dgab734

126. Ishigaki Y, Kawagishi N, Hasegawa Y, Sawada S, Katagiri H, Satomi S, et al. Liver Transplantation for Homozygous Familial Hypercholesterolemia. J Atheroscler Thromb 2019;26(2), 121–127. doi:10.5551/jat.RV17029

127. Martinez M, Brodlie S, Griesemer A, Kato T, Harren P, Gordon B, et al. Effects of Liver Transplantation on Lipids and Cardiovascular Disease in Children With Homozygous Familial Hypercholesterolemia. Am J Cardiol 2016;118(4):504–10. doi:10.1016/j.amjcard.2016.05.042

128. El-Rassi I, Chehab G, Saliba Z, Alawe A, Jebara V. Fatal cardiac atherosclerosis in a child 10 years after liver transplantation: a case report and a review. J Clin Lipidol 2011;5(4):329–32. doi:10.1016/j.jacl.2011.05.002

129. Elisofon SA, Magee JC, Ng VL, Horslen SP, Fioravanti V, Economides J, et al. Society of pediatric liver transplantation: Current registry status 2011-2018. Pediatr Transplant 2020;24(1):e13605. 10.1111/petr.13605

130. de Ville de Goyet J, Baumann U, Karam V, Adam R, Nadalin S, Heaton N, et al. European Liver Transplant Registry: Donor and transplant surgery aspects of 16,641 liver transplantations in children. Hepatology 2022;75(3):634–45. 10.1002/hep.32223

131. Baumann U, Karam V, Adam R, Fondevila C, Dhawan A, Sokal E, et al. Prognosis of Children Undergoing Liver Transplantation: A 30-Year European Study. Pediatrics 2022;150(4). doi:10.1542/peds.2022-057424

132. Rawal N, Yazigi N. Pediatric Liver Transplantation. Pediatr Clin North Am 2017;64(3):677–84. doi:10.1016/j.pcl.2017.02.003

133. Mansoorian M, Kazemi K, Nikeghbalian S, Shamsaeefar A, Mokhtari M, Dehghani SM, et al. Liver transplantation as a definitive treatment for familial hypercholesterolemia: A series of 36 cases. Pediatr Transplant 2015;19(6):605–11. doi:10.1111/petr.12562

134. Revell SP, Noble-Jamieson G, Johnston P, Rasmussen A, Jamieson N, Barnes ND. Liver transplantation for homozygous familial hypercholesterolaemia. Arch Dis Child. 1995;73(5):456–8. doi:10.1136/adc.73.5.456

135. Cephus CE, Qureshi AM, Sexson Tejtel SK, Alam M, Moodie DS. Coronary artery disease in a child with homozygous familial hypercholesterolemia: Regression after liver transplantation. J Clin Lipidol 2019;13(6):880–6. 10.1016/j.jacl.2019.09.007

136. Mlinaric M, Bratanic N, Dragos V, Skarlovnik A, Cevc M, Battelino T, Groselj U. Case Report: Liver Transplantation in Homozygous Familial Hypercholesterolemia (HoFH)—Long-Term Follow-Up of a Patient and Literature Review. Front Pediatr 2020;8:567895. doi: 10.3389/fped.2020.567895

137. Schmidt HH, Tietge UJ, Buettner J, Barg-Hock H, Offner G, Schweitzer S, et al. Liver transplantation in a subject with familial hypercholesterolemia carrying the homozygous p.W577R LDL-receptor gene mutation. Clin Transplant 2008;22(2):180–4. doi:10.1111/j.1399-0012.2007.00764.x

138. Al Dubayee M, Kayikcioglu M, van Lennep JR, Hergli N, Mata P. Is Liver Transplant Curative in Homozygous Familial Hypercholesterolemia? A Review of Nine Global Cases. Adv Ther 2022;39(6):3042–3057. doi:10.1007/s12325-022-02131-3

139. Barbir M, Khaghani A, Kehely A, Tan KC, Mitchell A, Thompson GR, Yacoub M. Normal levels of lipoproteins including lipoprotein(a) after liver-heart transplantation in a patient with homozygous familial hypercholesterolaemia. Q J Med 1992;85(307-308):807–12.

140. Greco M, Robinson JD, Eltayeb O, Benuck I. Progressive Aortic Stenosis in Homozygous Familial Hypercholesterolemia After Liver Transplant. Pediatrics 2016;138(5). doi:10.1542/peds.2016-0740

141. De Gucht V, Cromm K, Vogt A, Julius U, Hohenstein B, Spitthover RM, et al. Treatment-related and health-related quality of life in lipoprotein apheresis patients. J Clin Lipidol 2018;12(5):1225–33. doi:10.1016/j.jacl.2018.05.008

142. Kayikcioglu M, Kuman-Tunçel O, Pirildar S, Yílmaz M, Kaynar L, Aktan M, et al. Clinical management, psychosocial characteristics, and quality of life in patients with homozygous familial hypercholesterolemia undergoing LDL-apheresis in Turkey: Results of a nationwide survey (A-HIT1 registry). J Clin Lipidol 2019;13(3):455–67. doi:10.1016/j.jacl.2019.02.001

